# Genetic overlap between treatment-resistant schizophrenia and smoking initiation

**DOI:** 10.1101/2025.09.24.25336556

**Authors:** Elise Koch, Helin H. Mohammad, Nadine Parker, Hasan Ç. Lenk, Lars A. R. Ystaas, Piotr Jaholkowski, Alexey A. Shadrin, Oleksandr Frei, Anders M. Dale, Srdjan Djurovic, Espen Molden, Kevin S. O’Connell, Ole A. Andreassen

**Affiliations:** Centre for Precision Psychiatry, Division of Mental Health and Addiction, Oslo University Hospital, and Institute of Clinical Medicine, University of Oslo, Oslo, Norway; Department of Clinical and Molecular Medicine, Faculty of Medicine and Health Sciences, Norwegian University of Science and Technology, Trondheim, Norway; Center for Psychopharmacology, Diakonhjemmet Hospital, Oslo, Norway; KG Jebsen Centre for Neurodevelopmental Disorders, University of Oslo and Oslo University Hospital, Oslo, Norway; Center for Bioinformatics, Department of Informatics, University of Oslo, 0316 Oslo, Norway; Center for Multimodal Imaging and Genetics, J. Craig Venter Institute, La Jolla, CA, USA; University of California San Diego, La Jolla, CA, USA; Department of Medical Genetics, Oslo University Hospital and University of Oslo, Oslo, Norway

**Keywords:** Treatment-resistant schizophrenia, smoking, clozapine, genetics, GWAS

## Abstract

Early detection of treatment-resistant schizophrenia (TRS) is of substantial clinical importance. While TRS is heritable, the associated genetic variants have been difficult to identify. Cigarette smoking is associated with non-response to antipsychotics, and smoking behavior and schizophrenia have a shared genetic basis. Thus, TRS may also have a shared genetic basis with smoking behavior. Here we aim to identify genetic variants associated with TRS, by leveraging overlapping genetic variants with smoking initiation to increase statistical power. We analyzed genome-wide data for TRS and smoking initiation with the conditional/conjunctional false discovery rate (cond/conjFDR) to identify shared loci, and LD score regression to determine genetic correlations. To investigate potential causal effects of shared loci, we performed Mendelian randomization (MR) analyses. Shared loci were mapped to genes, which were further investigated for enrichment of drug target genes. We observed a significant positive genetic correlation between TRS and smoking initiation (r_g_ = 0.47 p = 0.0002). Leveraging the genetic overlap between TRS and smoking initiation, we identified four novel loci jointly associated with TRS and smoking initiation. The condFDR results improved polygenic prediction of TRS. MR showed putative evidence for a causal effect of genetic liability to TRS on smoking initiation. The functional genetic analyses showed that alpha-1-adrenergic receptors are likely involved in the pathophysiology of TRS and possibly related to the efficacy of clozapine versus other antipsychotic drugs. In conclusion, our results show that shared genetic mechanisms influence both TRS and smoking behavior, which provide new insights into the biological underpinnings of TRS.

## Introduction

Treatment-resistant schizophrenia (TRS) is defined as a failure of response to at least two antipsychotic drugs administered in adequate dose and duration, and occurs in about 30% of patients with schizophrenia^1^. TRS has been associated with increased mortality, higher rates of suicide attempts, and longer hospital stays, leading to substantial economic burdens due to healthcare utilization^2^. The antipsychotic drug clozapine is superior to other antipsychotics in terms of clinical effect and long-term outcomes^3-5^. However, due to the risk of rare but potentially fatal hematological adverse effects^5-7^, clozapine’s only indication is TRS. Clozapine treatment is effective in approximately 60% of TRS cases^8^, and may decrease mortality in schizophrenia^3, 9^. Thus, early identification of TRS is of substantial clinical importance, but a significant challenge is the high clinical and biological heterogeneity that characterizes TRS^10^.

Genome-wide association studies (GWAS) have identified hundreds of genetic loci harboring risk variants for schizophrenia^11^. While little progress has been made in identifying genetic associations with pharmacological treatment outcomes in psychiatry^12^, emerging evidence suggests that TRS may have a genetic component^13, 14^. The identification of genetic variants associated with TRS is limited by insufficient sample sizes as well as variability in definitions of TRS status^15^. No genome-wide significant loci have yet been identified in a GWAS of TRS defined based on the use of clozapine, including the world’s largest sample of 30,826 individuals with schizophrenia (N_TRS_ = 10,501 and N_non-TRS_ = 20,325)^13^.

The prevalence of cigarette smoking in schizophrenia is estimated to 70-80%, which is about three times the rate in the general population^16^. Hundreds of loci have been identified for tobacco use in a GWAS including over 1 million individuals^17^, where smoking phenotypes were positively genetically correlated with schizophrenia^17^. A Mendelian randomization study showed shown that genetic liability to cigarette smoking significantly increases the risk of developing schizophrenia^18^. However, the underlying factors driving the high smoking rates in schizophrenia patients are unclear^16^. It has been hypothesized that schizophrenia patients smoke to self-medicate or alleviate negative and cognitive symptoms^19, 20^. Furthermore, cigarette smoking has been associated with non-response to antipsychotics^21^. In TRS, cigarette smoking has been associated with more severe negative symptoms^22^. While it is known that smoking induces CYP1A2^23^, which metabolizes antipsychotic drugs such as clozapine and olanzapine^24, 25^, cigarette smoking also decreases the efficacy of olanzapine treatment independently of CYP1A2 genotype^26^. These findings indicate that smoking behavior and non-response to antipsychotics may have a shared genetic basis.

Considering the limited understanding of the underlying genetics of TRS based on the GWAS findings so far, complementary statistical approaches are needed to gain better understanding of the genetic underpinnings of TRS and to explore the genetic relationship between TRS and smoking behavior. Hypothesizing that TRS has a shared genetic basis with smoking behavior, we aim to identify genetic variants associated with TRS, by leveraging overlapping genetic associations with smoking initiation to increase power for genetic discovery.

## Methods

### GWAS sample description

We utilized publicly available GWAS summary statistics for TRS, which included 10,501 TRS cases and 20,325 non-TRS patients, with data derived from the CLOZUK and Psychiatric Genomics Consortium (PGC) cohorts^13^. For smoking initiation (“ever/never smoked regularly”), data were derived from a large-scale GWAS on smoking initiation in a population sample (N = 1,232,091)^17^. All GWAS data utilized in this study are from individuals of European ancestry. The Norwegian Institutional Review Board for the South-East Norway Region has evaluated the current protocol and found that no additional institutional review board approval was needed because no individual data were used.

### Conditional/conjunctional FDR and genetic correlation analysis

To boost discovery of genetic variants associated with TRS, we applied the conditional false discovery rate (condFDR) approach^27, 28^, using default settings (**Supplementary Methods**). The condFDR re-ranks genetic variants compared to p-value-based ranking and increases the power to discover loci associated with a primary phenotype (TRS) by leveraging associations with a secondary phenotype (smoking initiation)^27-29^. We then performed conjunctional FDR (conjFDR) analyses to identify shared variants between TRS and smoking initiation. In conjFDR, the process of the condFDR analysis is repeated switching the roles of the primary and secondary phenotypes. The largest condFDR value between the two runs is then used as the conjFDR value. A variant with a conjFDR less than 0.05 was considered as a shared variant (corresponding to 5 false positive per 100 reported associations). To estimate bivariate genetic correlations between TRS and smoking initiation, we utilized linkage disequilibrium score regression (LDSC)^30^.

### Locus definition, functional annotation and gene mapping

To define genetic loci based on the association summary statistics produced with conjFDR, we used FUMA^31^with default settings. Using FUMA^31^, each lead SNP per identified locus was annotated with Combined Annotation Dependent Depletion (CADD)^32^scores, which predict how deleterious the SNP effect is on protein structure/function, and RegulomeDB^33^scores, which predict the likelihood of regulatory functionality of SNPs. To investigate previous phenotype associations, the identified loci were queried in the GWAS catalogue^34^. Loci were also queried in the GTEx portal (GTEx v8)^35^for known expression quantitative trait loci (eQTLs) across multiple tissues. Identified lead variants were mapped to genes using the Open Targets Genetics platform (https://genetics.opentargets.org/)^36^that provides a Variant to Gene (V2G) association score for each variant-gene prediction, to assign likely causal genes for a given variant. For each lead variant, we considered the top three genes with the highest V2G score as well as the closest gene in case it was not among the top three genes with the highest V2G score. To identify drug-gene interactions, mapped genes were queried in the drug-gene interaction database (dgidb) v5.0.8 (06/12/2024)^37^. More details can be found in **Supplementary Methods**.

### Mendelian Randomization

To estimate the potential causal relationship of TRS on smoking initiation as well as of smoking initiation on TRS, we used Mendelian randomization (R version 4.1.1, TwoSampleMR version 0.5.6)^38^and reported results for inverse variance weighted^39^, weighted median^40^, weighted mode^41^, and MR Egger^42^(**Supplementary Methods**). Since TRS did not have any genome wide significant SNPs (p<5e-08), we used a reduced threshold for SNP inclusion (p<1e-05) when TRS was set as the exposure. This is consistent with previous Mendelian randomization studies with GWAS of low statistical power^43, 44^.

### Polygenic prediction of treatment-resistant schizophrenia

Using PRSice-2^45^, polygenic scores (PGSs) were calculated using summary statistics for TRS^13^and smoking initiation^46^. We compared PGS based on standard GWAS-ranked lead SNPs with PGS based on condFDR-based ranking, applying the pleioPGS approach^47^, using variant effect sizes derived from the original TRS GWAS. We calculated the PGSs for specific numbers of lead SNPs rather than for significance thresholds to allow for direct comparison between the approaches, at equal numbers of SNPs in each set. We compared the top 100 (approximate number of SNPs p<1E-5 in the original TRS GWAS) to 105,000 SNPs (number of independent SNPs p<1 in the original TRS GWAS). Sex, age and the first 10 genetic principal components were included as covariates.

The target sample includes 1,635 individuals (819 TRS and 816 non-TRS) from the therapeutic drug monitoring (TDM) service at the Center for Psychopharmacology in Diakonhjemmet Hospital, Oslo, Norway, between January 2005 and August 2022. TRS was defined based on the use/prescription of clozapine, the main drug indicated in TRS^48^, verified by detectable serum concentrations of clozapine. Non-TRS patients had no history of clozapine treatment and had only used other antipsychotics (according to TDM records). Of the 1,635 individuals, 912 were males (481 TRS and 431 non-TRS) and 723 were females (338 TRS and 385 non-TRS). The Regional Committee for Medical and Health Research Ethics and the Investigational Review Board at Diakonhjemmet Hospital approved the study. More information as well as information about genotyping and imputation can be found in **Supplementary Methods**.

### Genetically informed drug prioritization

Genes identified from Open Targets Genetics^36^were studied within networks of protein-protein interactions (PPIs) of gene products, using the latest version of the human protein interactome^49^, consisting of 18,217 unique proteins (nodes) interconnected by 329,506 PPIs after removing self-loops. As most approved drugs do not target disease-associated proteins but bind to proteins in their network vicinity^50^, we defined a network not only including the genes identified from Open Targets Genetics^36^, but also genes in their immediate network proximity. To define a TRS network, we used the method network propagation^51-53^, implemented in the Cytoscape^54^application Diffusion^53^. Genes identified from Open Targets Genetics^36^were used as input query genes, and the top 1% of proteins from the diffusion output were included in the TRS network. The Drug Gene Interaction Database (DGIdb, (http://www.dgidb.org/)v.5.0.8^37^was used to identify drug-gene interactions between approved drugs and genes in the TRS network. Gene-set enrichment analysis (GSEA) was performed to test for enrichment of drug-gene interactions within the TRS network (more details in **Supplementary Methods**).

## Results

### Genetic overlap between treatment-resistant schizophrenia and smoking initiation

Genetic correlation analyses showed a significant positive correlation between TRS and smoking initiation (r_g_= 0.4716, SE = 0.1277, z-score = 3.6921, p-value = 0.0002). The conditional QQ-plots indicate cross-trait polygenic enrichment between TRS and smoking initiation (**Figure S1**). This is demonstrated by the leftward deflection, showing an increase in associations with TRS as a function of significance in smoking initiation. At conjFDR < 0.05, we identified four loci jointly associated with TRS and smoking initiation (**Table 1, Figure 1**). The same four loci were identified at condFDR < 0.05, with the only difference that the locus on chromosome 16 had another lead SNP (rs9928337). A list of all candidate variants in the identified loci is provided in **Table S1**(condFDR) and **Table S2**(conjFDR). Investigation of the four identified loci in the GWAS catalog^34^showed that one locus (lead SNP rs494904) has been previously associated with both alcohol use disorder and problematic alcohol use^55^, while no associations have been reported for the other identified loci.

**Table 1:** Novel loci for treatment resistant schizophrenia (TRS) identified through conditional and conjunctional FDR analysis with smoking initiation.

| Lead SNP | Locus (chr: start-end) | A1/A2<sup>#</sup> | Mapped genes | p-value in TRS GWAS | Beta (SE) in TRS GWAS | p-value in Smoking GWAS | Beta (SE) in Smoking GWAS | conjFDR |
| --- | --- | --- | --- | --- | --- | --- | --- | --- |
| rs12030126 | 1: 236808558 - 236854973 | G/T | *LGALS8*, *HEATR1*, ***ACTN2*** | 3.51e-5 | -0.112 (0.027) | 2.14e-5 | 0.007 (0.002) | 0.01752 |
| rs494904 | 2: 45130410 - 45157336 | C/T | ***SIX3***, *SIX2* | 5.77e-5 | 0.100 (0.025) | 7.63e-11 | 0.010 (0.002) | 0.02762 |
| rs4076010 | 2: 137066601 - 137119376 | T/C | *DARS1*, ***CXCR4***, *MCM6* | 1.08e-4 | 0.090 (0.023) | 2.283-3 | 0.004 (0.002) | 0.04924 |
| rs9935028 (rs9928337) | 16: 25382373 - 25454780 | G/A | ***ZKSCAN2***, *AQP8*, *HS3ST4*, (*LCMT1*) | 3.56e-6 | -0.104 (0.022) | 9.07e-4 | 0.005 (0.002) | 0.00861 |

Lead SNPs in independent genomic loci jointly associated with TRS and smoking initiation at $\text{cond}/\text{conjFDR}$ threshold $< 0.05$ . Genomic regions separated by less than 250 kb were merged into a single locus. The table lists chromosomal positions (chr) and includes the top three mapped genes identified through Open Targets Genetics<sup>36</sup>. The nearest gene to the index SNP (in GRCh37 coordinates) is in bold font. The gene *LCMT1* was among the top three mapped genes to the lead SNP identified in $\text{condFDR}$ (rs9928337). <sup>#</sup>A1 is the effect allele and A2 is the reference allele.

**Figure 1.**
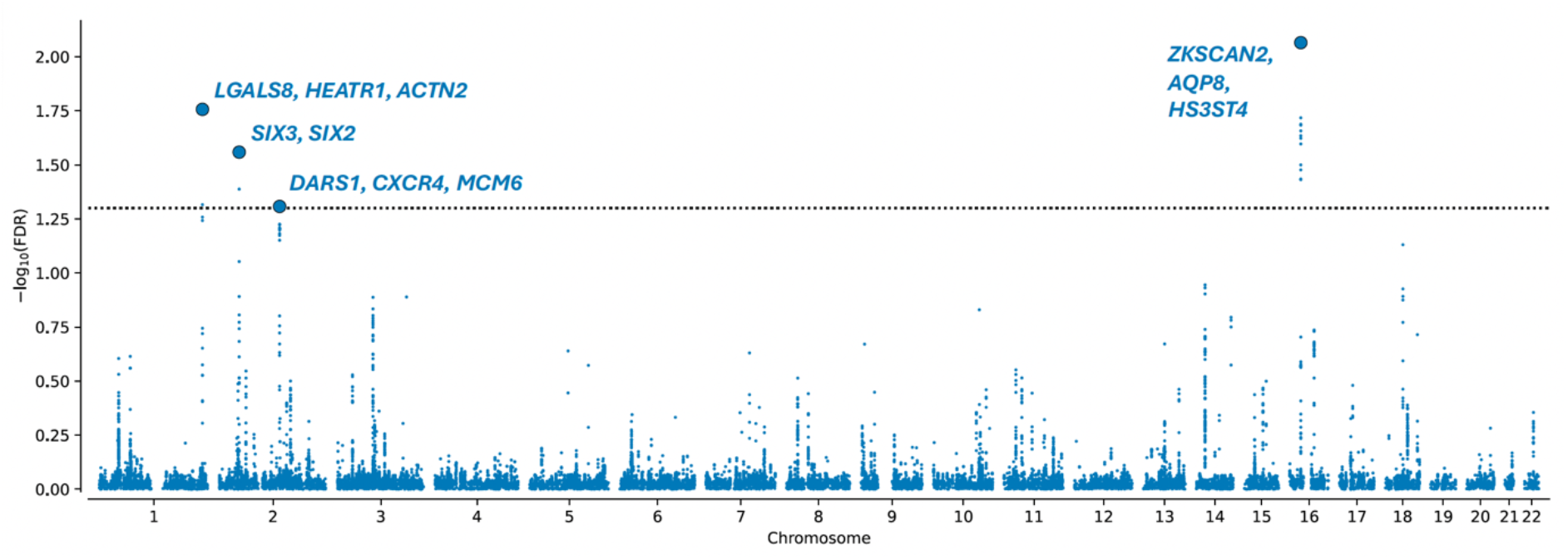
Manhattan plot showing genetic variants associated with TRS and smoking initiation by conjunctional FDR analysis, conjFDR < 0.05. The y-axis represents the -log10 transformed conjFDR values for each SNP, and the x-axis represents chromosomal positions. The dotted horizontal line indicates the threshold for significant shared associations (conjFDR < 0.05). Blue circles over the threshold represent lead SNPs with their respective top three genes with the highest variant-to-gene scores.

Functional annotations for these loci do not suggest the lead SNPs to be deleterious (CADD scores <12.37)^32^or likely to have regulatory functionality (RegulomeDB scores = 5-7)^33^. Assessment of the variant-gene relationships in the GTEx database^35^showed significant associations between the lead SNP rs12030126 on chromosome 1 and *LGALS8*gene expression, with most significant associations in adipose tissue (p=4.2e^-7^), the anterior cingulate cortex (p=1.7e^-5^), and nucleus accumbens (p=5.4e^-5^). The SNP was also associated with *HEATR1*gene expression, which was most significant in the esophagus (2.7e^-12^) and cerebellum (p= 6.6e^-11^), as well as with expression of *ACTN2*in esophagus (2.8e^-19^). The lead SNP rs9935028 on chromosome 16 was associated with expression of *ZKSCAN2*in muscle (p=2.5e^-9^) and adipose tissue (p=7.0e^-5^). Further assessment of the mapped genes in the DGIdb^37^showed that *AQP8*interacts with 7 approved drugs (**Table S3**), most of which are prostaglandin analogs or derivates indicated for the treatment of hypertension and/or hormonal or reproductive functions. Moreover, *CXCR4*interacts with 5 approved drugs primarily indicated for cancer treatment, and *ACTN2*interacts with adenosine triphosphate (**Table S3**). No interactions with approved drugs were identified for the other genes.

### Mendelian Randomization

Although the GWAS of TRS lacked power to estimate causal effects on smoking initiation using genome-wide significant loci, a relaxed threshold (p<1e-5) showed a putative causal link to smoking initiation (p<0.05) using the inverse variance weighted, weighted median, and weighted mode methods (**Table S4**). No evidence of a causal effect of smoking on TRS was seen (**Table S4**).

### Polygenic prediction of treatment-resistant schizophrenia

We compared the top 100-105,000 SNPs using original TRS GWAS p-value ranking and condFDR-based ranking (**Figure S1**), hypothesizing that the boosted power from our conditional analysis would select more informative variants than standard GWAS, resulting in improved PGS performance. The PGS based on condFDR-ranked SNPs explained more of the variance in TRS compared to both the standard TRS PGS and the smoking initiation PGS (**Figure S1**), with the highest variance explained (R^2^= 0.0079, p=0.002) at 52,000 SNPs. The highest variance explained by the TRS PGS was at 65,000 SNPs (R^2^=0.0065, p=0.005), while condFDR-based ranking achieved equivalent predictive performance by using almost half the number of SNPs (R^2^=0.0066 at 37,000 SNPs), which indicates a more efficient capture of relevant genetic signal with condFDR-based ranking. **Figure 2** shows the PGS performance in TRS for the PGS based on condFDR-ranked SNPs and the standard TRS PGS at 10,000, 20,000, 30,000, 40,000, 50,000, and 105,000 SNPs. The smoking initiation PGS is also plotted for comparison.

**Figure 2.**
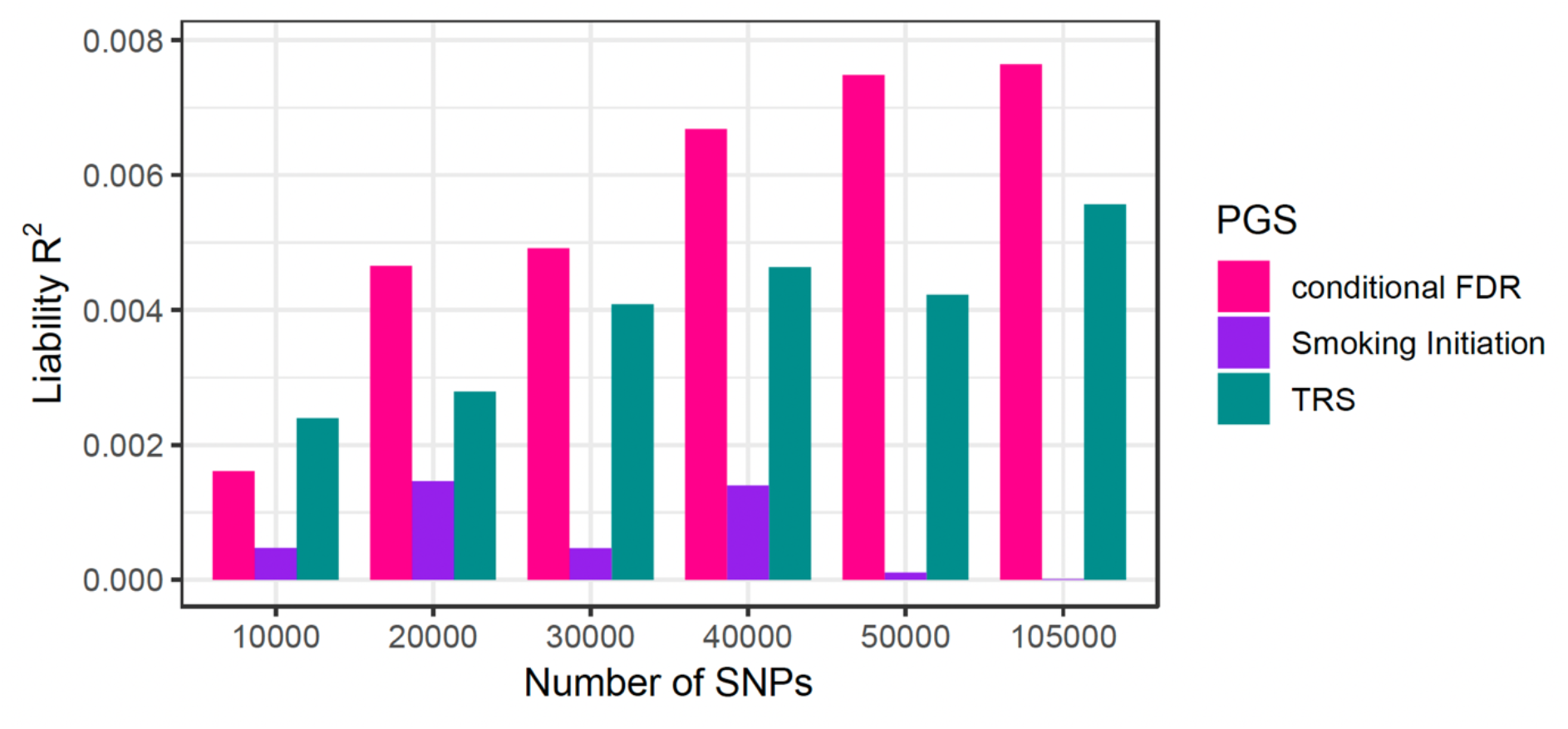
Explained variance (on the liability scale) in treatment-resistant schizophrenia (TRS) for polygenic scores (PGS) based on three different SNP rankings: TRS (green), smoking initiation (purple), and conditional FDR-based ranking of TRS conditioned on smoking initiation (pink). PGS were constructed for top 100 – 105,000 independent SNPs from the TRS GWAS summary statistics. The results of explained variance for all SNP ranges are presented in Figure S2.

### Genetically informed drug prioritization

The genes included in the TRS network (N=194) and the corresponding diffusion output values can be found in **Table S5**. Drug target genes in the TRS network were enriched (p<0.05) for targets of 11 drugs, most of which were alpha-1 adrenergic receptor agonist/antagonist, used to manage cardiovascular conditions. However, after correcting for the total number of drug-gene interactions (N=25,290), none of the enrichments remained significant (FDR>0.05) (**Table S6**). The TRS network and the drugs identified based on gene-set enrichment analyses are shown in **Figure 3**.

**Figure 3:**
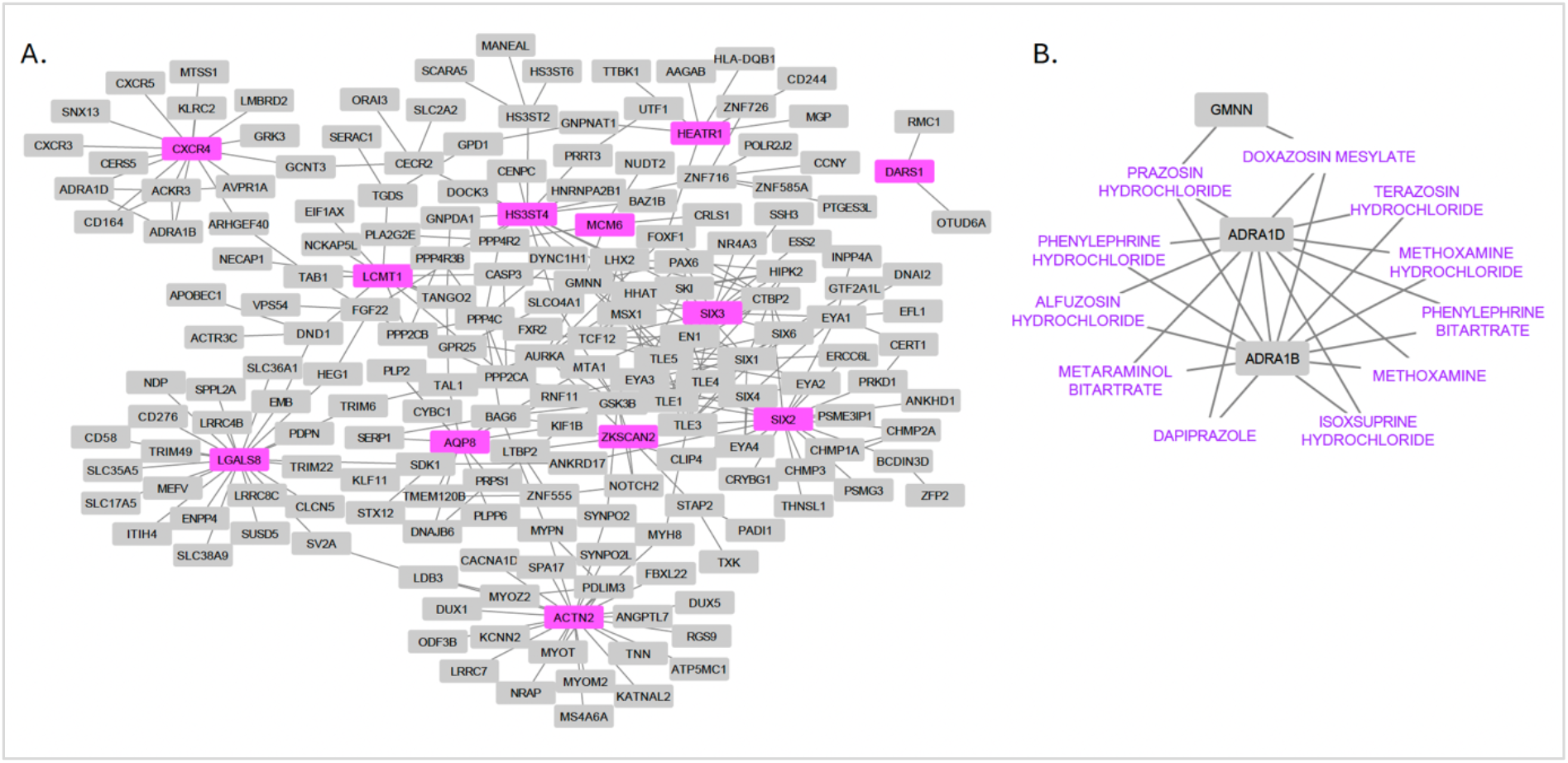
Treatment-resistant schizophrenia network (**A**) and drug target genes in the network that were enriched for alpha-1 adrenergic receptor modulators (**B**). The input genes to build the network are highlighted in pink. Nodes refer to genes or drugs, and edges refer to gene-drug interactions or gene-gene interactions through identified protein-protein interactions between gene products (proteins).

## Discussion

In the present study, we boosted discovery of genetic variants associated with TRS and identified four novel loci associated with TRS after conditioning on smoking initiation. Genetic correlation analysis showed a positive genetic correlation between TRS and smoking initiation, and Mendelian randomization analyses showed putative evidence for a causal effect of TRS on smoking initiation. In the independent validation sample, a PGS based on condFDR-ranked SNPs showed greater variance explained in TRS compared to the standard TRS PGS. From PPI network-based analyses and GSEA, we identified alpha-1-adrenergic receptors as potential targets for TRS.

Applying the condFDR framework^27-29^, we increased discovery in an underpowered GWAS by leveraging genetic overlap with a second, well-powered GWAS^56, 57^. We identified four novel loci for TRS, while no loci were identified in the original TRS GWAS^13^. Some of the mapped genes have been previously associated with schizophrenia. *LGALS8*(mapped to rs12030126 on chromosome 1), encoding Galectin-8, is downregulated in the hippocampus of schizophrenia patients^58^. It has also been linked to cigarette smoking, with evidence suggesting that cigarette smoke-induced autophagy impairment leads to increased galectin-8 and inflammation^59^. While *ZKSCAN2*(mapped to the rs9935028 on chromosome 16) has not been previously associated with schizophrenia, other members of the zinc finger transcription factors family (e.g., ZKSCAN3 and ZKSCAN4) have shown associations with gray matter reduction in schizophrenia^60, 61^. Aquaporins such as AQP3 and AQP4 have been associated with the pathophysiology of schizophrenia^62, 63^. While AQP8 (mapped to rs9935028 on chromosome 16) is expressed in the brain, its function in the CNS remains unclear ^64^. Alpha-actinin-2, encoded by *ACTN2*(mapped to the locus on chromosome 1, rs12030126), could indirectly associate with schizophrenia and possibly TRS through its role in interacting with glutamate N-methyl-D-aspartate (NMDA) receptors^65^, which have been linked to the pathophysiology of both schizophrenia and TRS^66^. The chemokine receptor type 4 (*CXCR4*, mapped to rs4076010 on chromosome 2) has also shown interactions with NMDA receptors, leading to the reduced NMDA receptor signaling that has been associated with the pathophysiology of schizophrenia^67^. Finally, mutations in the *SIX3*gene (mapped to rs494904 on chromosome 2) have been associated with both schizophrenia^68^and smoking^69^.

Several studies have shown significant genetic correlations between schizophrenia and smoking initiation (r_g_ranging between 0.10 and 0.16)^17, 70, 71^, indicating a shared genetic basis. In a GWAS of smoking behaviors among schizophrenia cases, it was demonstrated that a PGS for smoking initiation was partially shared between schizophrenia cases and the general population^70^. However, the molecular mechanisms underlying the schizophrenia-smoking association are not completely understood^70^. While a genetic overlap between schizophrenia and smoking behavior has been demonstrated in studies applying the cond/conj FDR method^72, 73^, it remains unknown if shared genetic factors with smoking initiation are different among TRS and non-TRS patients. We demonstrate a significant positive genetic correlation between TRS and smoking initiation that is significantly higher than the genetic correlation between smoking initiation and schizophrenia, supporting a shared genetic basis between TRS and smoking initiation that is independent from the genetic contribution of schizophrenia. By leveraging the identified genetic overlap between TRS and smoking initiation, we show that a
PGS based on condFDR-ranked SNPs explained the greatest variance in TRS compared to both the standard TRS PGS and the smoking initiation PGS, albeit the predicted value was small. This pleioPGS approach outperformed the standard GWAS-based ranking, utilizing less SNPs in the PGS, despite using the same SNP weightings, indicating a more efficient capture of relevant genetic signal with condFDR-based ranking.

The network analyses implicated two alpha-1-adrenergic receptors (ADRA1B and ADRA1D), and we shortlist several drugs acting on these receptors. Results from several clinical trials have demonstrated that alpha-1-adrenergic receptor antagonists are efficacious in the treatment of negative symptoms and social functioning deficits in schizophrenia^74-76^. Clozapine, which shows superior efficacy compared to conventional antipsychotic drugs, shows significant affinity for alpha-adrenergic receptors, especially alpha-1-adrenergic receptors^77^. Of note, clozapine exerts an advantageous therapeutic effect on negative and cognitive symptoms, which are usually rather resistant to treatment with other antipsychotics^78^. Because clozapine displays significant affinities for several neurotransmitter receptors including muscarinic, histaminergic, and adrenergic receptors, with comparatively low D_2_dopamine receptor binding^77, 79^, a critical question is which of these receptor affinities may contribute to clozapine’s superior therapeutic effect^79, 80^. It has been hypothesized that clozapine’s superior efficacy is related to its alpha-adrenoceptor modulation, stating that alpha-1- and alpha-2-adrenoceptor blocking stabilizes the dysregulated central dopaminergic systems in schizophrenia^80^. Although the mechanisms involved remain to be fully understood, it has been suggested that antagonism of alpha-1-adrenergic receptors may suppress striatal hyperdopaminergia involved in positive symptoms, while alpha-2-adrenergic receptor antagonism may improve prefrontal dopaminergic functioning thereby reducing negative and cognitive symptoms^80^. Moreover, alpha-1-adrenoceptor antagonism has been related to the metabolic side effects of antipsychotics^77, 81^, with clozapine being associated with the largest degree of metabolic dysfunction^82^. Interestingly, it has been suggested that clozapine’s effectiveness may be related to its metabolic side effects^56, 82^. Taken together, these results indicate that alpha-adrenergic receptor antagonism may be considered as treatment for TRS after further investigation.

It should be noted that the cond/conjFDR method does not identify the specific causal variants underlying the overlapping genomic loci, and that the detection of cross-trait enrichment is influenced by the power of the investigated GWAS^29^. Although our results highlight how data from low-powered GWASs can still be useful to study genetic architecture and overlap between traits, our results might be influenced by the small sample size of the TRS GWAS. However, despite identifying novel loci for TRS, larger GWAS of TRS are required to better understand the underlying genetics of TRS. In addition, the predictive ability of the PGS remains far from being clinically relevant, and larger GWAS of TRS will also improve SNP weights for polygenic prediction. The phenotyping of the TRS GWAS samples have not recorded comorbid smoking in the schizophrenia cases, and there could potentially be differences in smoking frequency between TRS and non-TRS patients. Moreover, the smoking GWAS samples may include schizophrenia cases, which could suggest that some of the genetic overlap may be due to shared phenotypes. In addition, nicotine and clozapine may have overlapping pharmacological mechanisms of action on acetylcholine receptors, which may be captured in the identified genetic overlap between smoking initiation and TRS defined by clozapine use. Finally, the individuals included in the GWAS used in our cond/conjFDR analyses were of predominantly European ancestry, which suggest that the findings may not translate to other ancestry groups.

In conclusion, by conditioning on smoking initiation, we boosted discovery and identified four novel loci associated with TRS. These findings suggest that shared genetic mechanisms influence TRS and smoking behavior. In addition, the results indicate that alpha-1-adrenergic receptors are involved in the pathobiology of TRS and are likely being related to the superior efficacy of clozapine. Together, our results provide new insights into the biological underpinnings of TRS.

## Data Availability

All data produced in the present study are available upon reasonable request to the authors.

## Author contributions

EK, KSO, and OAA conceived the study and were involved in study design. EK, HHM, and NP conducted analyses. EK drafted the initial manuscript. All authors contributed to data interpretation and editing of the manuscript.

## Acknowledgements

We thank the research participants, employees, and researchers of the CLOZUK study, PGC, GSCAN, Center for Precision Psychiatry, University of Oslo, Norway, and Center for Psychopharmacology, Diakonhjemmet Hospital, Oslo, Norway for making this research possible.

## Funding

This work was partly performed on the TSD (Tjeneste for Sensitive Data) facilities, owned by the University of Oslo, operated and developed by the TSD service group at the University of Oslo, IT-Department (USIT). Computations were also performed on resources provided by UNINETT Sigma2—the National Infrastructure for High Performance Computing and Data Storage in Norway (NS9666S). We gratefully acknowledge support from the Research Council of Norway (RCN) (296030, 223273, 334920, 300309, 326813, 324252). This project has received funding from the European Union’s Horizon 2020 research and innovation programme under grant agreement No 964874 (REALMENT), and from the European Union’s Horizon 2021 research project (EU HORIZON-HLTH-2021, grant 101057454).

## Conflict of Interest

Dr. Andreassen reported grants from Stiftelsen Kristian Gerhard Jebsen, South-East Regional Health Authority, Research Council of Norway, and European Union’s Horizon 2020 during the conduct of the study; personal fees from cortechs.ai (stock options), Lundbeck (speaker’s honorarium), and Sunovion (speaker’s honorarium) and Janssen (speaker’s honorarium) outside the submitted work. Dr. Anders M. Dale is Founding Director, holds equity in CorTechs Labs, Inc. (DBA Cortechs.ai), and serves on its Board of Directors. Dr. Dale is the President of J. Craig Venter Institute (JCVI) and is a member of the Board of Trustees of JCVI. He is an unpaid consultant for Oslo University Hospital. All other authors report no financial interests or potential conflicts of interest.

## Code and data availability

The code for cond/conjFDR and pleioPGS is publicly available at https://github.com/precimed/pleiofdrand https://github.com/norment/open-science/tree/main/2021_VanderMeer_medRxiv_pleioPGS, respectively.

## Notes

### Competing Interest Statement

The authors have declared no competing interest.

