## Supplementary_materials for "Genetic overlap between treatment-resistant schizophrenia and smoking initiation"

### Supplementary Figures

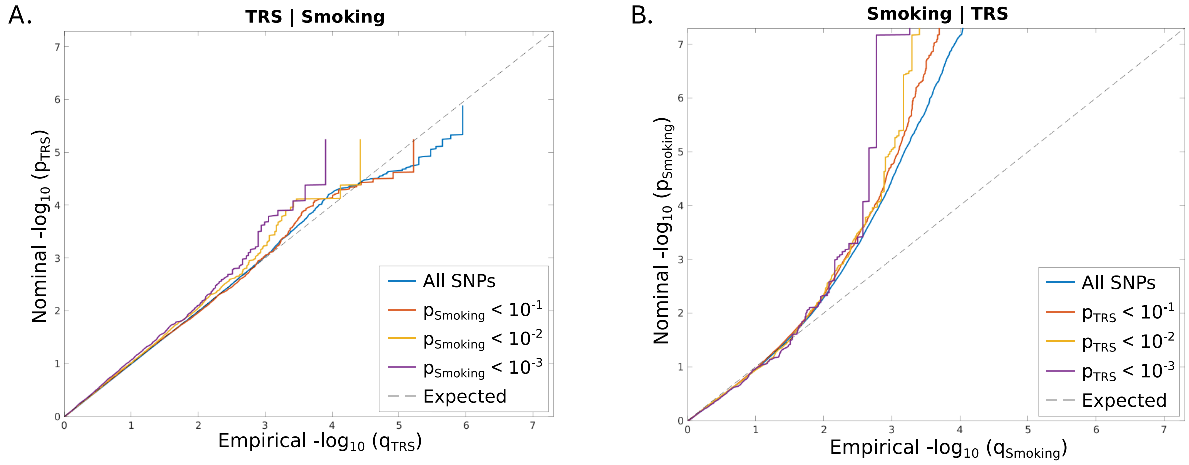


**Figure S1**. Conditional QQ-plots. (**A**): Conditional QQ-plot showing cross-phenotype polygenic enrichment between treatment-resistant schizophrenia (TRS) and smoking initiation. Plotted are the nominal vs empirical −log10 p-values (corrected for inflation) for TRS, below the standard genome-wide association study threshold of p < 5e−8, as a function of significance of association with smoking initiation at the levels of p < 0.10, p < 0.01, and p < 0.001. (**B**): Conditional QQ-plot showing cross-phenotype polygenic enrichment between smoking initiation and TRS. Plotted are the nominal vs empirical −log10 p-values (corrected for inflation) for smoking initiation, below the standard genome-wide association study threshold of p < 5e−8, as a function of significance of association with TRS at the levels of p < 0.10, p < 0.01, and p < 0.001. The dashed line indicates the null hypothesis.

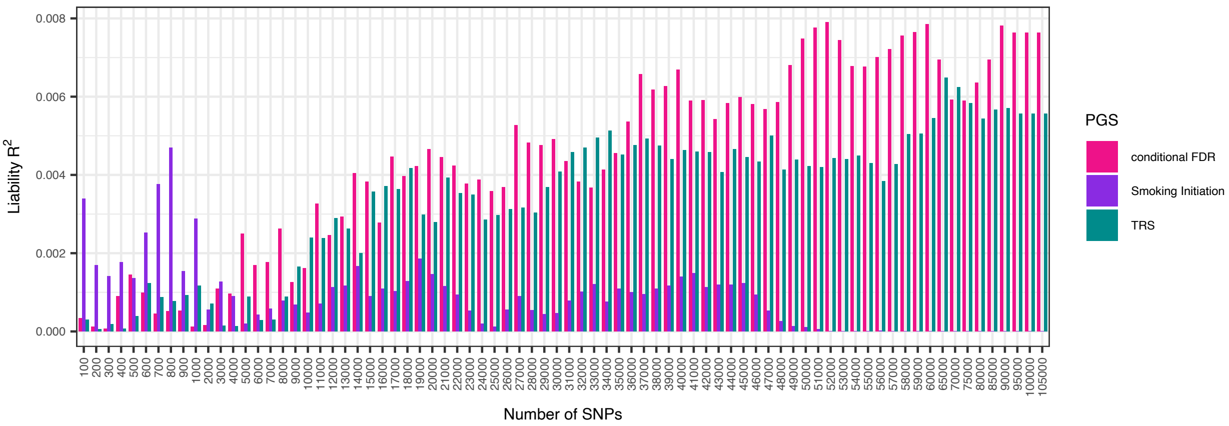


**Figure S2**. Explained variance (on the liability scale) in treatment-resistant schizophrenia for polygenic scores (PGS) based on three different SNP rankings: Treatment-resistant schizophrenia (TRS, green), smoking initiation (purple), and conditional FDR-based ranking of TRS conditioned on smoking initiation (pink). PGS were constructed for top 100 – 105,000 independent SNPs from the TRS GWAS summary statistics.

### Supplementary Methods

#### *Conditional/conjunctional false discovery rate*

To assess the cross-phenotype polygenic enrichment between treatment-resistant schizophrenia (TRS) and smoking initiation, we constructed conditional QQ-plots by comparing enrichment of association in all variants in the primary trait (TRS) with three nested subsets of variants defined by their strength of association ($p < 0.1$, $p < 0.01$ and $p < 0.001$) with the secondary trait (smoking initiation). A successive left-ward deflection of the QQ-plot with increasing significance indicates cross-trait enrichment and can be directly interpreted according to the Bayesian definition of the true discovery rate (TDR = 1-FDR), whereby a larger shift is consistent with a smaller FDR. The condFDR estimates a posterior probability that a single nucleotide polymorphism (SNP) has no association in the primary phenotype, given that p-values of the SNP in both the primary and conditional phenotypes are lower than observed p-values. Genomic control procedure utilizing intergenic variants was applied to correct GWAS p-values prior to condFDR analyses[1]. Analyses were repeated 100 times with randomly pruned sets of SNPs. For each iteration, a single SNP in each linkage disequilibrium (LD)-independent region ($r^2$ >0.1) was retained, the results were then averaged across all iterations. The European sample of the 1000 Genomes Project phase 3[2] was used as LD reference, in PLINK version 1.9[3]. SNPs within the extended major histocompatibility complex (MHC) region and chromosome 8p23.1 (genome build hg19 locations chr6:25119106–33854733 and chr8:7200000–12500000, respectively) were excluded when fitting the condFDR model, as the high LD of these regions may bias condFDR estimation[4].

***Locus definition, variant annotation, and gene mapping***

To define genetic loci based on the association summary statistics produced with conjFDR, we used FUMA[5] with default settings. Genetic variants with conjFDR < 0.05 and with an LD $r^2$ < 0.6 with each other were defined as independent significant variants. Of these, variants with an LD $r^2$ < 0.1 were selected as lead variants. For a given lead variant the borders of the genomic locus were defined as minimum/maximum positional coordinates over all corresponding candidate variants. Loci that were separated by less than 250 kb were then merged. Using FUMA[5], SNPs were annotated with Combined Annotation Dependent Depletion (CADD)[6] scores, which predict how deleterious the SNP effect is on protein structure/function, and RegulomeDB[7] scores, which predict the likelihood of regulatory functionality of SNPs. To investigate previous phenotype associations, the identified loci were queried in the GWAS catalogue[8]. Loci were also queried in the GTEx portal (GTEx v8)[9] for known expression quantitative trait loci (eQTLs) across multiple tissues. Identified lead variants were mapped to genes using the Open Targets Genetics platform (https://genetics.opentargets.org/)[10]. This portal contains functional genomics data from different cell types and tissues retrieved from various repositories and datasets, including data on gene expression, chromatin conformation, and protein abundance that are aggregated to make robust connections between variants and likely causal genes. For each variant-gene prediction, a Variant to Gene (V2G) association score is provided to assign likely causal genes for a given variant. For each locus, we considered the top three genes with the highest V2G score as well as the closest gene in case it was not among the top three genes with the highest V2G score.

***Mendelian Randomization***

Mendelian randomization (MR) is an approach that estimates causal associations between two phenotypes (i.e., exposure and outcome) by leveraging the fact that individuals are randomly assigned genetic risk for the exposure of interest[11]. Genetic variants associated with the exposure phenotype are selected as instruments for MR analyses. The MR framework is based on three major assumptions (i) the instruments are associated with the exposure, (ii) the instruments are not associated with confounders, (iii) the instruments influence the outcome only through the risk factor. It is important to note that violations of these assumptions produce unreliable MR estimates. Therefore, it has become general practice to use multiple robust methods of MR to test causal associations. There are several methods to perform MR. One of the more robust approaches is two-sample MR which uses GWAS summary statistics, from independent samples, for the exposure and outcome phenotypes of interest. Here, we performed two-sample MR to investigate the causal relationship between TRS (exposure) and smoking initiation (outcome). We also performed MR to investigate the causal relationship between smoking initiation (exposure) and TRS (outcome). For the MR analyses, we used the summary statistics from a GWAS on TRS[12], and a GWAS on smoking initiation[13]. These analyses were performed to investigate if genetic liability to TRS has a causal effect on smoking initiation and if genetic liability to smoking initiation has a causal effect on TRS. For smoking initiation, we included SNPs at a level of p<5e-8, while we lowered the p-value threshold for TRS to p<1e-5 (as no genome-wide significant loci were identified in the TRS GWAS). We used the R package TwoSampleMR where we report results using the following methods: inverse variance weighted[14], weighted median[15], weighted mode[16], and MR Egger[17]. The inverse variance weighted approach is a classic MR method which is essentially a fixed-effect meta-analysis of the SNP effects on the outcome over the SNP effects on the exposure. For increased validity of our MR outcomes, we incorporated sensitivity analysis techniques (the weighted median, weighted mode, and MR Egger approach) in addition to the inverse variance weighted approach. These approaches are more robust to heterogeneity in estimates (i.e., a potential sign of violation of MR assumptions) than the inverse variance weighted approach.

***Norwegian Therapeutic Drug Monitoring sample***

The study used anonymized data and residual blood samples from already performed routine analyses. In these cases, where study inclusion does not pose any burden to the patients, the Health Research Act of Norway allows for using samples and information collecting in clinical routine for research purpose without written informed patient consent. However, it is mandatory to send information to the patients, in advance of starting a project, about their legal rights to reserve against being included. Accordingly, letters were sent to all patients eligible for inclusion, where those requesting to opt-out were excluded from the study. DNA was extracted from whole blood, and genotyping was performed with the Human Omni Express-24 v.1.1 array (Illumina Inc., San Diego, CA, USA) in accordance with the standard Illumina protocol, at deCODE Genetics (Reykjavik, Iceland). Standard pre-imputation quality control was performed using PLINK v1.9[3, 18]. We used the TOPMed imputation server (https://imputation.biodatacatalyst.nhlbi.nih.gov/#!) to impute genetic variants in the full cohort. Briefly, the quality control parameters for retaining SNPs and subjects were: SNP missingness <0.05 (before sample removal), subject missingness <0.02, SNP missingness <0.02 (after sample removal), SNP Hardy–Weinberg equilibrium (p-value $<1 \times 10^{-6}$). Furthermore, related individuals (pairwise Identity-By-Descent >0.2 according to PLINK v1.9[3, 18]) were removed.

***Genetically informed drug prioritization***

To define a TRS network, we used the method network propagation[19-21], implemented in the Cytoscape[22] application Diffusion[21]. Starting with a chosen set of input proteins, information from their PPIs is transferred to all other proteins in the interactome and received from them through an iterative process. Network proximity between proteins is scored depending on their PPIs, where higher diffusion output values relate to higher relatedness to the input proteins[19-21]. Genes identified from Open Targets Genetics[10] were used as input query genes, and the top 1% of proteins from the diffusion output were included in the TRS network.

The Drug Gene Interaction Database (DGIdb, (https://www.dgidb.org/) v.5.0.8 (06/12/2024)[23] was used to identify drug-gene interactions between approved drugs and genes in the TRS network. The DGIdb provides information on drug-gene interactions from 28 diverse sources that are aggregated and normalized. The database collects drug-gene interactions based on information about therapeutic targets and their corresponding drugs, knowledge from clinical trials, as well as potentially clinically actionable drug-gene associations based on metadata such as molecule structure and molecular weight[23]. To prioritize drugs targeting genes in our TRS network, the genes in the network were tested for enrichment for drug-gene interactions. This was performed using gene set enrichment analyses (GSEA) applying hypergeometric testing and gene sets (drug target genes) provided by the DGIdb[23], to identify enrichment of genes targeted by specific drugs. For each drug, an enrichment score was calculated as the ratio between drug-gene interactions in the network genes and all drug-gene interactions in the DGIdb[23].
